# Demethylation and upregulation of an oncogene post hypomethylating treatment

**DOI:** 10.1101/2020.07.21.20157776

**Authors:** Yao-Chung Liu, Junsu Kwon, Emiliano Fabiani, Yanjing V. Liu, Chong Gao, Giulia Falconi, Lia Valentini, Carmelo Gurnari, Adrianna I. Jones, Junyu Yang, Henry Yang, Julie A. I. Thoms, Ashwin Unnikrishnan, John E. Pimanda, Rongqing Pan, Maria Teresa Voso, Daniel G. Tenen, Li Chai

**Affiliations:** Department of Pathology, Brigham and Women’s Hospital, Boston, MA 02115, USA; Division of Hematology, Department of Medicine, Taipei Veterans General Hospital, Taipei, Taiwan; Faculty of Medicine, School of Medicine, National Yang-Ming University, Taipei, Taiwan; Program in Molecular Medicine, School of Life Science, National Yang-Ming University, Taipei, Taiwan; Cancer Science Institute of Singapore, National University of Singapore, 117599, Singapore; Department of Biomedicine and Prevention, University of Tor Vergata, Rome, Italy; Harvard Stem Cell Institute, Harvard Medical School, Boston, MA 02115 USA; School of Medical Sciences and Lowy Cancer Research Centre, Faculty of Medicine, UNSW Sydney, NSW 2052, Australia; Prince of Wales Clinical School and Lowy Cancer Research Centre, Faculty of Medicine, UNSW Sydney, NSW 2052, Australia; Department of Haematology, Prince of Wales Hospital, Randwick, NSW 2031, Australia; Department of Medical Oncology, Dana-Farber Cancer Institute, Boston, MA, 02115

## Abstract

**Background:** While hypomethylating agents (HMA) are currently used to treat myelodysplastic syndrome (MDS) and patients with cancer, their effects on reactivation and/or upregulation of oncogenes are generally not well elucidated. SALL4 is a known oncogene that plays an important role in MDS. In this study, we examined the impact of HMA on SALL4 methylation and expression.

**Methods:** Paired bone marrow samples from a cohort of MDS patients on the BMT-AZA trial, collected before and after four cycles of azacytidine (AZA) treatment, were used to explore the relationship between changes in SALL4 expression, treatment response and clinical outcome with a follow-up of up to 40 months. No/low-SALL4 expressing leukemic cell lines were used to study the relationship between SALL4 methylation and expression. A novel locus-specific demethylation technology, CRISPR-DNMT1-interacting RNA (CRISPR-DiR), was used to identify the CpG island critical for SALL4 expression.

**Results:** In MDS patients, we noted SALL4 upregulation after AZA treatment in 40% of the cases. Significantly, patients with SALL4 upregulation had a worse outcome. Using CRISPR-DiR, we discovered that demethylation of a 500bp CpG island within the 5’UTR-Exon1-Intron1 region was critical for SALL4 expression. Importantly, in cell lines and patients, we confirmed that HMA treatment led to demethylation of the same CpG region and upregulation of SALL4 expression.

**Conclusions:** CRISPR-DiR was useful to define the critical region important for gene activation. Along with analysis of patient samples, we demonstrated that demethylation and upregulation of an oncogene after HMA treatment can indeed occur and should be further studied.

## INTRODUCTION

Myelodysplastic syndrome (MDS) comprises heterogeneous myeloid disorders characterized by cytopenias and dysplasia in peripheral blood and bone marrow (BM), with ineffective hematopoiesis and a variable risk of leukemic transformation.^1,2^ The revised International Prognostic Scoring System (IPSS-R) provides a reliable estimation of the risks of leukemic transformation, progression, and death.^3,4^ In addition, MDS is characterized by high genetic heterogeneitywith rare cases bearing a single gene mutation, and often with a combination of mutations insplicing factors (e.g., *SF3B1, SRSF2, U2AF1, ZRSR2*; seen in approximately 40–60% of patients) and/or epigenetic regulators (e.g., *TET2, ASXL1, DNMT3A, EZH2, IDH1, IDH2*; observed in approximately 40–50%) in most patients.^5^

For high-risk (HR) patients, those that are elderly or unfit to receive chemotherapy, hypomethylating agent (HMA) therapy is the first-line treatment, and also used as a bridge to transplant in younger patients.^6,7^ The optimal timing to assess cytidine nucleoside analog azacytidine (AZA) response is after four to six cycles of treatment, and overall survival (OS) was reported to be prolonged in two thirds of patients receiving at least four cycles of treatment.^7,8^ Although continuous AZA therapy in responders was reported to be beneficial to improve patients’ clinical parameters, the survival after AZA in ‘real-world’

HR-MDS/low-blast count acute myeloid leukemia (AML) was lower than the expected OS in clinical trials,^7^ and the outcome after AZA failure was less than 6 months.^9^ To date, there is no treatment available to improve OS after HMA failure. Therefore, the key is to identify biologic or molecular predictor(s) that could guide the clinician to decide on the timing of HMA-based combination regimens, which will hopefully improve clinical outcomes in patients with first-line HMA monotherapy.

Despite the known mode of action of the drugs, there seems to be little correlation between the degree of demethylation following HMA and hematologic response.^10^ We hypothesize that global HMA treatment not only contributes to the demethylation of tumor suppressor genes, but can also induce demethylation of oncogenes. In this study, we used the known oncogene SALL4 as an example to examine the effects of HMA on upregulation of oncogenes, and its association with treatment response and overall outcomes.

Spalt-like transcription factor 4 (SALL4) plays an essential role in MDS and AML leukemogenesis^11,12^ and tumorigenesis in various solid tumors.^13-15^ SALL4 is reactivated or aberrantly expressed in a multitude of cancers; and identified by meta-analysis as a promising poor prognostic factor implicated in cancer recurrence.^16^ In hematological malignancies, SALL4 is aberrantly expressed in HR-MDS^17^, AML^12,18^, blast phase of chronic myeloid leukemia,^19^ and precursor B-cell lymphoblastic leukemia/lymphoma.^20^ In a murine model with constitutive SALL4 expression, mice developed MDS□like features and subsequently leukemic transformation through activation of the Wnt/beta-catenin pathway.^21^ These mice also demonstrated impairmentof DNA damage repair linked to the Fanconi anemia pathway.^22^ In MDS patients, the levels of SALL4 were high in HR-IPSS. Notably, the expression level of SALL4 significantly increased in MDS/AML transformation and decreased following effective therapy.^17^ Moreover, aberrant hypomethylation of SALL4 at diagnosis was more frequentlyfound in HR-MDS patients (14% in Low/Int-1 versus 39% in Int-2/High), and the survival of the SALL4-hypomethylated group was shorter than that of *SALL4*-methylated group.^23^ However, the effect of HMA on SALL4 expression and its clinical implications for MDS/AML patients are unknown.

Therefore, we retrospectively analyzed SALL4 expression and clinical survival of 25 MDS patients with BM samples before and after 4 cycles of AZA treatment, from a cohort of 37 patients enrolled in the BMT-AZA trial. To define the mechanism (s) of SALL4 modulation in response to AZA treatment, using a novel CRISPR-DNMT1-interacting RNA (CRISPR-DiR) approach, we first identified and demethylated the CpG island critical for SALL4 expression. We then evaluated the effects of HMA treatment on the methylation status of SALL4 in MDS patient samples.

## METHODS

### Patients and sample collection

BM samples were obtained from 37 newly diagnosed MDS patients enrolled in the BMT-AZA trial.^24,25^ Study was overseen and approved from the University of Tor Vergata, Rome.Written informed consent was received from participants prior to inclusion in the study. CD34^-^ (n = 10) and CD34^+^ (n = 5) BM mononuclear cells (BM-MNCs) from healthy donors were used as control cohorts. In 37 MDS patients, 25 patients had paired BM samples collected before and after 4 AZA cycles. BM-MNCs were isolated by Ficoll gradient centrifugation using Lympholyte-H (Cedarlane, Ontario, Canada), according to the manufacturer’s instructions. MDS diagnoses were according to the 2008 World Health Organization (WHO) classification.^26^ All 25 studied patients received at least 4 cycles of AZA treatment. Other clinical characteristics including age at diagnosis, sex, IPSS, WHO-Classification Based Prognostic Scoring System (WPSS), response after 4 cycles of AZA treatment, peripheral blood counts, BM blasts, and cytogenetics were also reviewed. The definition of first response follows the International Working Group (IWG) criteria.^27,28^ In the study, responders included patients achieving complete remission (CR), partial remission (PR) and hematologic improvement (HI), whereas nonresponders included those patients with stable disease (SD) and progressive disease (PD).

### Cell culture and treatment protocol

The K562 and HL-60 human leukemia cell lines were purchased from ATCC and cultured in RPMI 1640 supplemented with 10% fetal bovine serum and antibiotics (penicillin-streptomycin 100U/100 μg/mL) and maintained in an incubator with 5% CO2 atmosphere at 37°C. Based on previous publications ^29,30^, 100nM, 250nM, and 500nM 5-aza-2’-deoxycytidine (decitabine, DAC) daily was used to treat K562 and HL-60 cells for five days. Before adding new DAC or DMSO treatment, cells were washed with ice-cold PBS at 24-hour intervals. Cells were collected on day 5 for measuring SALL4 levels using droplet digital polymerase chain reaction (ddPCR) for RNA and western blot analysis for protein, as well as methylation studies.

Additional materials are described in the supplementary materials.

## RESULTS

### HMA treatment leads to upregulation of SALL4 and is associated with poor outcome

To evaluate the impact of HMA on SALL4 expression after treatment, we first measured the baseline SALL4 expression at diagnosis from BM-MNCs of MDS patients prior to AZA treatment. In 37 MDS patients, 33 were categorized as high/intermediate-2 riskaccording to IPSS, whereas 2 patients were intermediate-1/low risk. The risk of the remaining 2 patients was unknown due to unavailable cytogenetics. Levels of SALL4 mRNA were significantly higher in 37 MDS patients (*p* = 0.002) compared to that of healthy donors (Fig.S1A), similar to what has been reported previously.^18^Among the 37 enrolled MDS patients, there were 25 patients with available paired BM samples at diagnosis and after 4 cycles of AZA. Of these 25 patients, 48% were defined as responders, with 36% achieving CR, 8% PR, and 4% HI, respectively (Fig. 1A). There was no difference at baseline SALL4 expression at diagnosis prior AZA treatment among these groups (Fig. S1A). These 25 patients were then stratified according to the fold change in SALL4 mRNA expression pre- and post-AZA treatment.A waterfall plot depicting fold changes in SALL4 mRNA levels demonstrated that patients can be separated into two distinct groups (Fig.1B). Ten out of the twenty-five patients (40%) had ≥ 2-fold increase in SALL4 expression (SALL4^up^) whereas fifteen patients (60%) had ≥ 2-fold decrease in SALL4 expression (SALL4^down^) after 4 cycles of AZA treatment.

**Figure 1.**
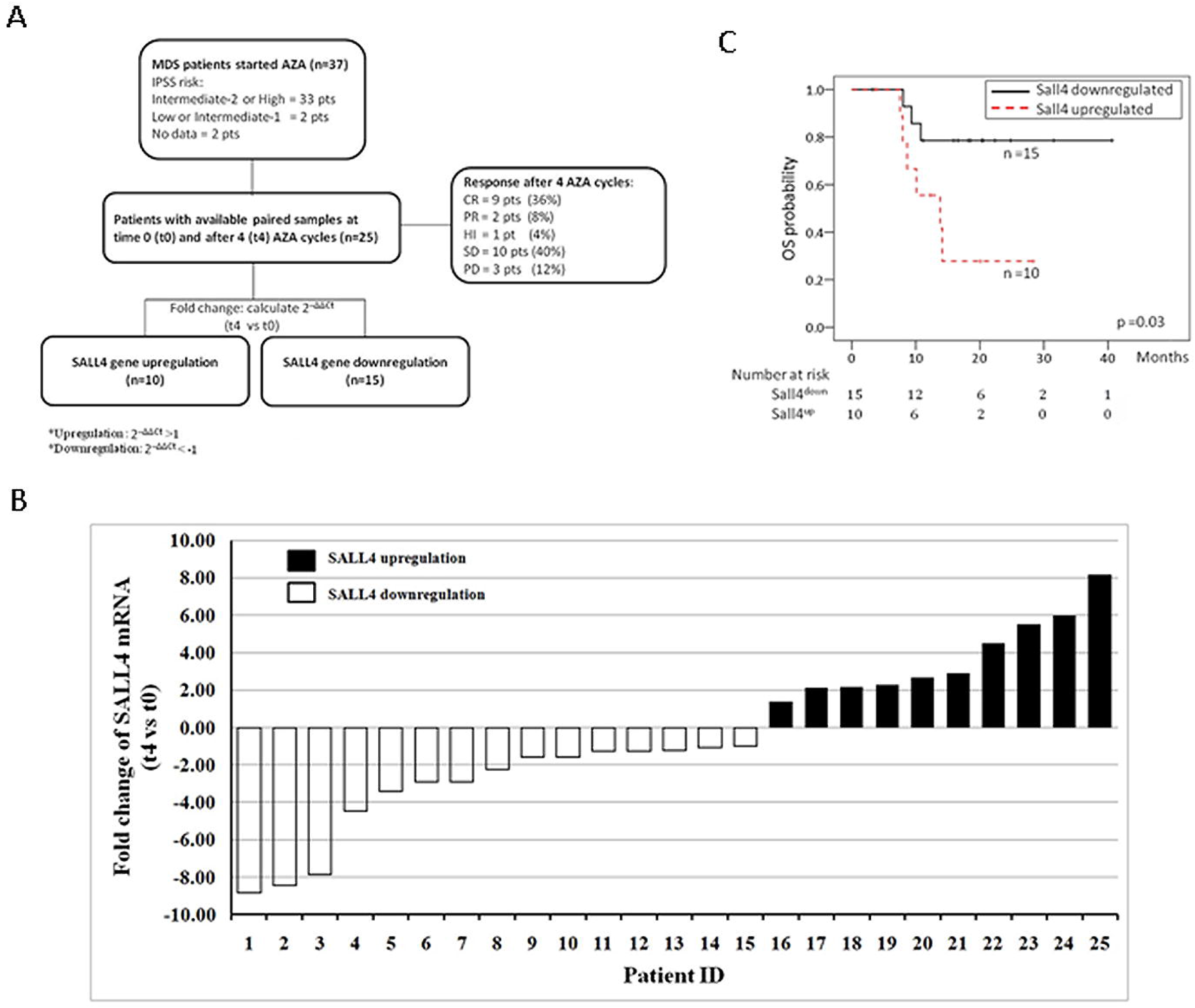
HMA treatment in MDS patients leads to increased SALL4 expression. (A) Flow chart of study design. (B) Waterfall plot of log_2_ fold change of SALL4 in 25 patients after 4 cycles of AZA treatment;expression changes in SALL4 mRNA from 25 patients with paired BM-MNC samples before and after 4 cycles of AZA treatment were measured by qRT-PCR. Ten out of the 25 patients (40%) had ≥ 2-fold increase in SALL4 expression (SALL4^up^) whereas 15 patients (60%) had ≥ 2-fold decrease in SALL4 expression (SALL4^down^) after four cycles of AZA treatment. The median log_2_ fold change for SALL4^up^ was 2.78 (IQR: 2.15□5.65), whereas SALL4^down^ was **□**2.25 with IQR from □1.26 to □4.45.(C) Overall survival (OS) between SALL4^up^ and SALL4^down^.

The median log_2_ fold change for SALL4^up^ was 2.78 (IQR: 2.15-5.65), whereas SALL4^down^ was −2.25 with interquartile ranges (IQR) from −1.26 to −4.45. There was no significant difference in fold change of SALL4 mRNA between responders and non-responders (Fig. S1B).

Intriguingly, when compared between the SALL4^up^ (n = 10) and SALL4^down^ (n = 15) groups, there was a trend towards a difference in progression□free survival (PFS) (*p* = 0.09, Fig. S2A) and a significant OS difference (*p* = 0.03, median: 13.8 months in the SALL4 upregulated vs. not reached in the SALL4 downregulated, Fig. 1C), indicating that SALL4 upregulation may be associated with worse survival.

Furthermore, there was significantly better PFS and OS in the responder/SALL4^down^ group than in the other three subgroups (*p* = 0.03 in PFS; *p* = 0.04 in OS). While the median survival in PFS or OS was not reached in the responder/SALL4^down^ subgroup, strikingly, in the responder/SALL4^up^ group, the median survival in PFS and OS was a dismal 7.9 months and 8.0 months, respectively (Fig. S2B & S2C).

The demographic and clinical-biological characteristics between the SALL4^up^ and the SALL4^down^patients are described in Table 1. In both groups, approximately 70% of patients subsequently received allogeneic stem cell transplantation. The median follow-up was 14.1 months (IQR: 9.0–20.4) after starting AZA. We further conducted Cox proportional-hazards model analysis for prognostic factors on OS based on characteristics at diagnosis, mutational profile^25^ (Fig. S3) and SALL4 expression changes. In multivariate analysis for OS, only SALL4^up^ (HR 6.48; 95% CI, 1.06 to 39.67; *p* = 0.04) and *RUNX1* mutation (HR 10.66; 95% CI, 1.25 to 90.72; *p* = 0.03) were independent negative predictors for OS (Table S1).

**Table 1.** Baseline characteristics of patients with SALL4 upregulation or downregulation after 4 cycles of azacytidine

| Characteristics | SALL4 upregulation | SALL4 downregulation | *p*-value |
| --- | --- | --- | --- |
|  | n=10 | n=15 |  |
| Median age,years (IQR) | 59.6 (46.2-61) | 59.5 (45-61.5) | 0.40 |
| Sex (%), male/female | 90/10 | 53.3/46.7 | 0.05 |
| IPSS risk (%) |  |  | 0.80 |
| Low or Intermediate-1 | 10 | 6.7 |  |
| Intermediate-2 or high | 90 | 86.7 |  |
| WPSS risk (%) |  |  | 0.50 |
| Very low or Low | 10 | 0 |  |
| Intermediate | 10 | 13.3 |  |
| High or very high | 80 | 66.7 |  |
| Responders/Nonresponders (%) | 50/50 | 46.7/53.3 | 0.87 |
| WBC (/μl), median (IQR) | 2450 (1900-4015) | 3320 (2280-10410) | 0.29 |
| ANC (/μl), median (IQR) | 400 (215-1490) | 700 (400-2000) | 0.33 |
| Hb (g/dL), median (IQR) | 9.9 (8.6-10.6) | 9.4 (8.7-11.5) | 0.40 |
| PLT count (/μl), median (IQR) | 49500 (26750-164250) | 77000 (49000-129000) | 0.46 |
| Median blasts in BM aspiration (%) | 13 (11-15) | 12 (3.8-17.8) | 0.39 |
| Cytogenetics for MDS (%)<sup>§</sup> |  |  | 0.82 |
| Good karyotype | 40 | 33.3 |  |
| Intermediate karyotype | 20 | 13.3 |  |
| Poor karyotype | 30 | 40 |  |
| Transplantation (%) | 70 | 73.3 | 0.85 |
| Mutational profiling (%)* |  |  |  |
| TET2 mutation | 10 | 40 | 0.17 |
| ASXL1 mutation | 30 | 40 | 0.69 |
| RUNX1 mutation | 20 | 26.7 | 0.70 |
| SETBP1 mutation | 20 | 26.7 | 0.70 |
| TP53 mutation | 30 | 13.3 | 0.35 |
| ZRSF2 mutation | 40 | 6.7 | 0.12 |
| DNMT3A mutation | 10 | 20 | 0.62 |
| SRSF2 mutation | 10 | 20 | 0.62 |
| SF3B1 mutation | 10 | 13.3 | 0.80 |
| **U2AF1 mutation** | **30** | **0** | **0.05** |
| CEBPA mutation | 0 | 20 | 0.25 |
| CBL mutation | 10 | 6.7 | 0.76 |
| ETV6 mutation | 0 | 13.3 | 0.50 |
| IDH2 mutation | 10 | 6.7 | 0.76 |
| NRAS mutation | 0 | 13.3 | 0.50 |
| CSF3R mutation | 10 | 0 | - |
| JAK2 mutation | 10 | 0 | - |

IQR, Interquartile range; SALL4, Spalt like transcription factor 4; IPSS, International Prognostic Scoring System; WPSS, WHO-Classification Based Prognostic Scoring System; WBC, White blood cell; ANC, Absolute neutrophil count; Hb, hemoglobin; PLT, platelet; BM, Bone marrow; MDS, myelodysplastic syndrome
\*No mutation of BRAF, CALR, EZH2, FLT3, HRAS, IDH1, ABL1, KIT, KRAS, MPL, NPM1, PTPN11, and WT1 in both groups
§Three patients had no metaphase for cytogenetic analysis

### Demethylation of a critical CpG region leads to upregulation of SALL4

We next investigated the mechanism(s) of SALL4 up regulation post HMA treatment. One possibility is that HMA treatment can lead to demethylation of a CpG island critical for SALL4 expression. The differential methylation of the SALL4 locus has been observed in K562-induced pluripotency reprogrammed cells^31^. The major CpG island of SALL4 is located across the 2,000-bp locus including the 5’UTR-Exon 1-Intron 1region. Within this CpG island, methylation of 5’UTR-Exon 1-Intron 1, a specific 500bp DNA segment, was further observed to be negatively correlated with SALL4 expression in hepatocellular carcinoma (HCC) patients and cell lines.^32^ We further hypothesized that demethylation of this region could lead to upregulation of SALL4.

In order tolocalize the critical CpG region that responsible for regulation of SALL4 expression, and to test the causal relationship between DNA methylation of SALL4 RNA expression, we applied the CRISPR-DNMT1-interacting RNA (CRISPR-DiR) technique^33^, a novel approach of inducing locus-specific demethylation by blocking DNMT1 activity ^34^ in cell lines with no/low SALL4 expression.

In the CRISPR-DiR method, two loops of the guide RNA which are not required for guide function have been replaced with sequences which specifically interact with and inhibit DNA methyltransferase 1 (DNMT1).^33,34^ This modified guide RNA can achieve site-specific demethylation.^33^ We tested several site-specific guide RNAs around the major CpG island of SALL4, and only sgSALL4_1 (named as DiR_SALL4 here), targeting around the CpG 16 region, demonstrated effective demethylation ability.^32^ We further monitored the methylation of the 5’UTR-Exon 1-Intron 1 CpG region in HL-60 cells (no/low SALL4 expression) following treatment with CRISPR-DiR (Fig. 2A). Upon transduction of HL-60 cells with DiR_SALL4, significant demethylation changes were observed after 8 days (Fig.2B) as well as increased SALL4 transcript levels (Fig.2C) compared to a control scrambled gRNA (DiR-NT). Similar demethylation results by DiR_SALL4 wereobserved in other low/no SALL4 expressing SNU387 cells, while CRISPR-DiRs targeting neighbouring regions could not demethylate.^32^ This demonstrates that demethylation of this region by SALL4 locus specific CRISPR-DiR can lead to upregulation of SALL4.

**Figure 2.**
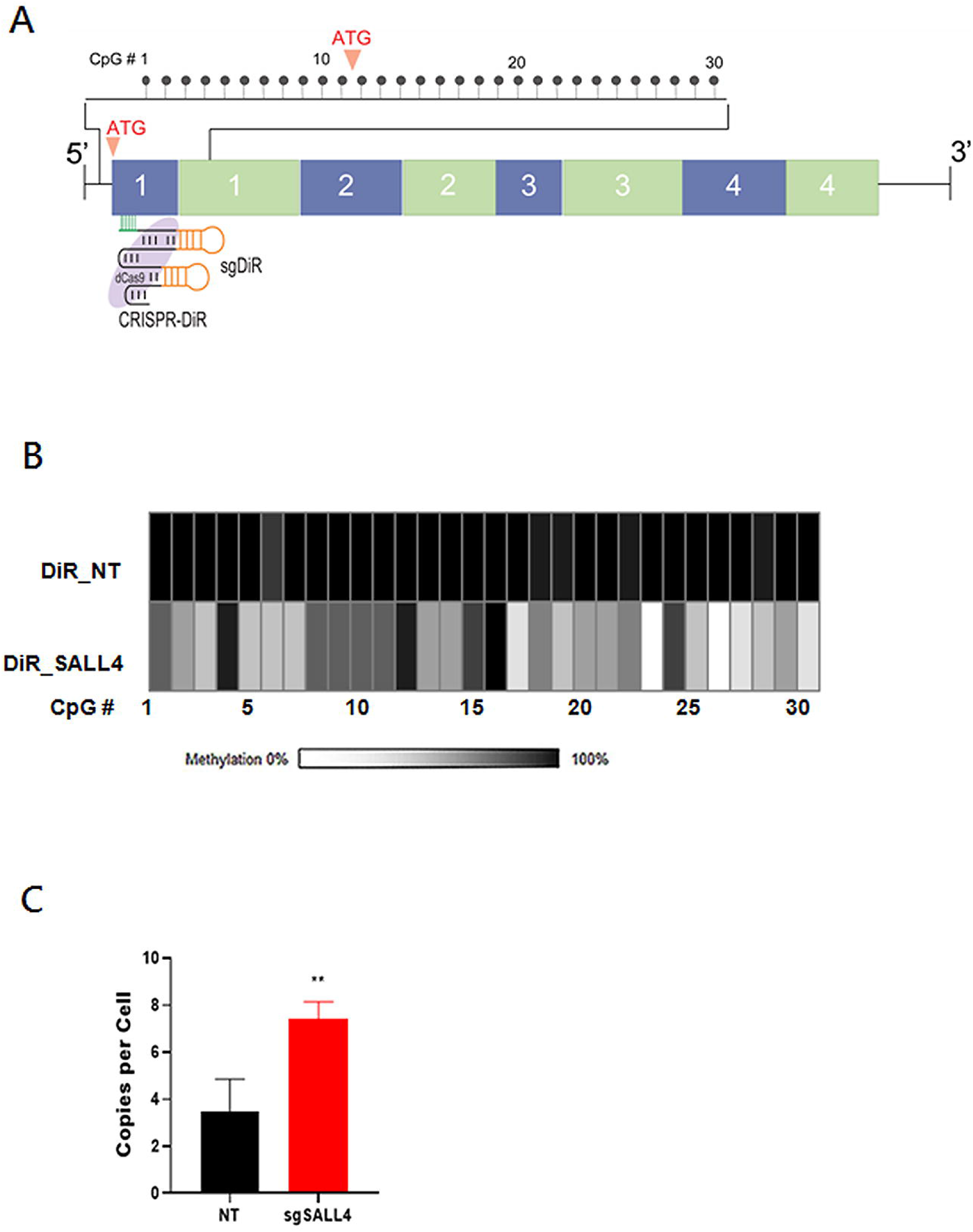
Demethylation of a critical CpG island and SALL4 expression in leukemic cells treated with CRISPR-DiR. (A) Schematic of the CpG region within the SALL45’ UTR – exon1– intron1 region. Depicted at the top of the diagram are the 30 CpG residues; below are depicted the 4 SALL4 exons (in blue and introns (in yellow), demonstrating the location of the 30 CpG residues in the exon1-intron1 region. The location of the target of the guide RNA for CRISPR-DiR is shown below, targeting CpG #16 and #17. (B) Bisulfite sequencing after CRISPR-DiR of HL-60 cells transduced with either a non-targeting negative control guide RNA (DiR_NT) or a targeting guide RNA (DiR_SALL4), leading to demethylation, with higher methylation indicated by darker shading. (C) SALL4 transcript (copies per cell assessed by ddPCR) upregulation in HL-60 cells treated with CRISPR-DiR 8 days after induction of the CRISPR-DiR.

### HMA treatment can lead to demethylation of SALL4

We then tested whether SALL4 could be upregulated by HMAs. Using HL-60 cells, we first evaluated the dynamics of SALL4 mRNA levels through a cycle of the HMA agent 5-aza-2’-deoxycytidine (DAC), using a dosage range commonly used in cell lines, from 100 nM to 500 nM^30^. The number of mRNA copies per cell (by ddPCR) and protein (by western blot) for SALL4 was measured at day 5. We noticed a dose dependent upregulation of SALL4 mRNA expression at day 5 (Fig. 3A and B).

**Figure 3.**
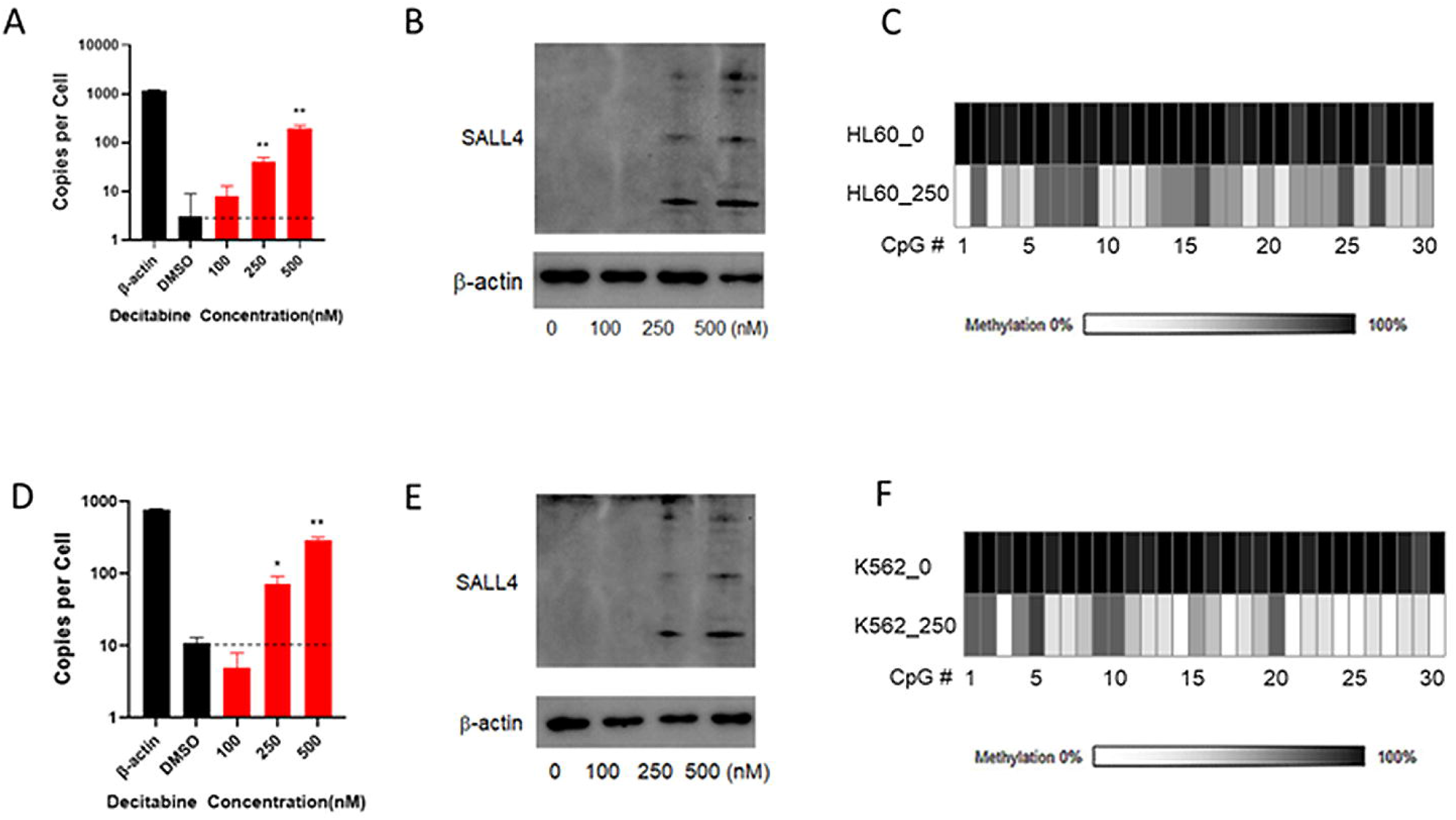
Demethylation of a critical CpG island and SALL4 expression in leukemic cells treated with HMA. (A) SALL4 copies per cell by ddPCR in HL-60 cells treated with DAC; (B) Western blot in HL-60 cells treated with DAC; (C) Methylation profiling in HL-60 cells untreated (HL60_NT) versus treated with 250 nM DAC (HL60_250); (D) SALL4 transcripts (copies per cell assessed by ddPCR) in K562 cells treated with DAC; (E) Western blot in K562 cells treated with DAC; (F) Methylation profiling in K562 cells treated with DAC. DAC treatment for all panels was for 5 days.

We next examined the methylation status of the SALL4 5’UTR-Exon 1-Intron 1 CpG island region after 250nM DAC treatment, and found this region was demethylated in HL-60 cells, in accordance with our CRISPR-DiR result (Fig. 3C). Similar results were also observed in another SALL4-low leukemic cell line, K562 (Fig. 3 D-F).

We next examined the methylation changes of MDS patient BM samples before and after HMA treatment. In four patients with increased SALL4 expression (SALL4^up^), we noticed decreased methylation at the critical CpG region (Fig. 4A&B). Conversely, we did not observe changes in the methylation at the same CpG region in three patients with decreased SALL4 expression (SALL4^down^) post-AZA treatment (Fig. 4C&D).

**Figure 4.**
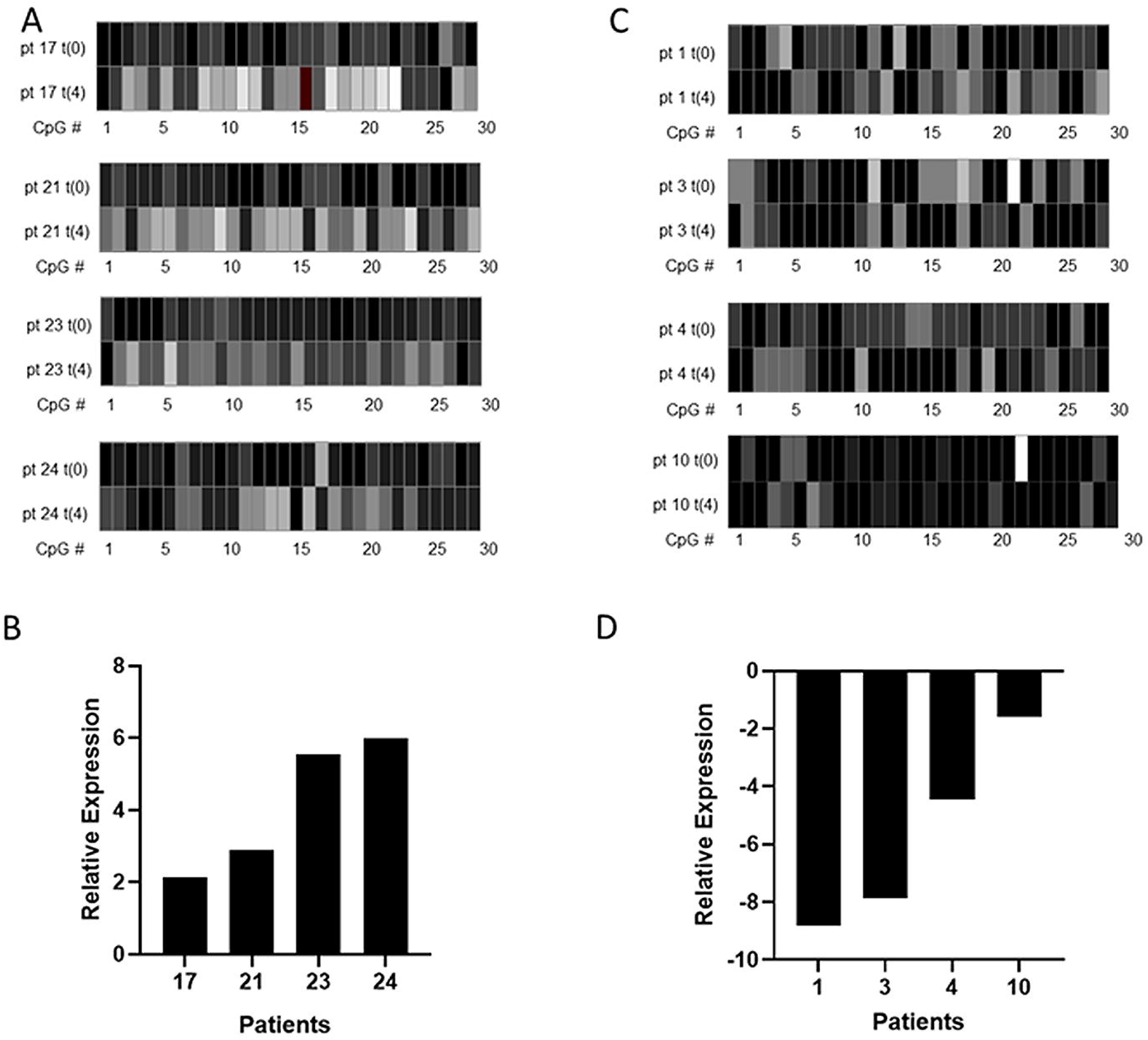
HMA treatment leads to demethylation of SALL4 in MDS patients. (A) Methylation changes after 4 cycles of AZA treatment in 4 patients in which SALL4 RNA is upregulated (Panel B). (C) Methylation changes after 4 cycles of AZA treatment in 4 patients in which SALL4 RNA is downregulated (Panel D); t(0) is baseline, t(4) is after 4 cycles of treatment.

Since genetic mutations, including epigenetic factors such as DNMT3A, IDH1, IDH2, and TET2 are common in MDS patients, we questioned whether the up or down regulation of SALL4 in patients post-AZA treatment was related to their pre-existing mutation status. Common somatic mutations in 30 genes, known to be frequently mutated in MDS patients, are reported below. At the diagnosis of MDS, we identified at least one mutation with more than 1% VAF in 22 out of 25 patients (88%). The median number of mutated genes was three per patient (range: 0-6). The most mutated genes in this population were: *ASXL1* (36%), *TET2* (28%), *RUNX1* (24%), *SETBP1* (24%), *ZRSF2* (20%), *TP53* (20%), *SRSF2* (16%), and *DNMT3A* (16%). Ten out of 22 patients had more than one gene mutation (Fig. S3). It is notable that U2AF1 mutations are only observed in the SALL4^up^ group (30%, 3 out of 10 patients, Table 1) and not in the SALL4^down^ group (0, 0 out of 10 patients). In addition, in SALL4^down^ group, NRAS, ETV6, and CEBPA mutations are observed, none of these gene mutations are detected in the SALL4^up^ group (Table 1).

## Discussion

SALL4 is an oncogene that plays an important role in MDS and AML.^11,12,17,18,22^ The level of SALL4 was reported as a molecular biomarker for diagnosis and prognosis of AML and for diagnosis of MDS.^11,12,21^ High levels of SALL4 transcripts and hypomethylation of *SALL4* were also significantly found in patients with higher IPSS or WPSS risk.^17^ Hypomethylation was recognized as a probable mechanism for aberrant high *SALL4* expression in MDS patients, but the effect of HMA on SALL4 expression has not been reported yet.

Using a novel CRISPR-DiR approach, we have identified a critical CpG island responsible for SALL4 expression, and HMA treatment can lead demethylation of this region and upregulation of SALL4. In our cohort of MDS patients (n = 25), we observed that after four cycles of AZA treatment, 40% of our patients (n = 10) were SALL4^up^ while 60% (n = 15) were SALL4^down^. While the fold change of SALL4 expression was not significantly different between clinical responders and non-responders, it was surprising to note the strikingly poor outcome of responders who have upregulated SALL4 (Fig. S2).In general, the responder group of MDS patients are considered to have a good outcome based on their clinical parameters after HMA treatment. However, the results from our study suggests that additional molecular biomarkers such as SALL4 may need to be monitored after HMA monotherapy in MDS patients. It is unclear to us why only a sub-set MDS patients has upregulation of SALL4 at the end of 4 cycles of AZA treatment, and future studies are necessary to reveal the underlying mechanism (s). A recent study by Reilly et al ^35^ suggested that certain genetic mutations can be grouped and associated with distinct DNA methylation (DNAm) cluster patterns.In our current study, we have noticed that the splicing factor U2AF1 mutations are only present in SALL4^up^group (Table 1). In Reilly’s study, U2AF1 mutations are enriched in DNAm cluster B and C, which are correlated with worse prognosis. U2AF1 mutation could affect DNMT3B expression.^36^ It is possible, when working with other mutations, U2AF1 mutations could contribute to establish a certain global DNA methylation stage, which in turn determine the long-term effects of HMA treatment on gene expression such as upregulation of SALL4.

Overall, our study suggests that HMA monotherapy can activate or upregulate oncogenes. In the case of SALL4, if SALL4 is upregulated, clinicians probably should not only rely on clinical parameters to predict the outcome of their patients. Instead, we propose that MDS patients on HMA therapy should be monitored for SALL4 expression in addition to traditional clinical parameters. While the upregulation of SALL4 may be associated with worrisome prognosis, it may also provide an additional treatment option based a SALL4 mediated cancer vulnerability. We are currently testing the ideas that if SALL4 expression is upregulated, whether alternative therapy that directly or indirectly mitigates SALL4 expression should be added to the treatment plan.^37^

## Data Availability

All data in the main manuscript and supplementary data are available on request

## Acknowledgements

This study was supported by AIRC 5×1000 call “Metastatic disease: the key unmet need in oncology” to MYNERVA project, #21267 (Myeloid Neoplasms Research Venture Airc). A detailed description of the MYNERVA project is available at http://www.progettoagimm.it. This work was also by Singapore Ministry of Health’s National Medical Research Council (Singapore Translational Research (STaR) Investigator Award, D.G.T.; NMRC/OFIRG/0064/2017.); Singapore Ministry of Education under its Research Centres of Excellence initiative; NIH/NCI Grant R35CA197697 and NIH/NHLBI P01HL131477 (D.G.T); as well as NIH/NHLBI Grant P01HL095489 (L.C.); Xiu research fund (L.C); National Health and Medical Research Council of Australia (JEP: APP1024364, APP1043934, APP1102589; AU: APP1163815) and Leukemia & Lymphoma Society USA Translational Program Grant 6589-20 (AU); and The Ministry of Science and Technology, Taiwan (MOST 109-2314-B-075-078 to Y.C.L).

