## Supplemental files for "Demethylation and upregulation of an oncogene post hypomethylating treatment"

**Affiliations:**

**Supplementary Methods**

***Statistics and study endpoints***

Clinical characteristics and mutational profiling are presented as the total number (*n*) and proportion (%). Data are presented as medians and interquartile ranges (IQR) for skewed data. The Fisher exact test or chi-squared test, as appropriate, were used to compare categorical variables, whereas continuous variables were compared using the Wilcoxon rank-sum test. Fold change of SALL4 mRNA levels between responders and non-responders was compared using the Mann-Whitney U test. Progression-free survival (PFS) was defined as the time from AZA treatment to disease progression or death from the treatment, whereas OS was defined as time from AZA treatment to death. OS and PFS were analysed using the Kaplan–Meier product–limit method with censoring for the patients who did not progress or die during the treatment period. A log–rank test was used to compare survival curves for statistical significance. Hazard ratios (HRs) and the 95% confidence interval (CI) were calculated using Cox proportional-hazards model. All prognostic factors with *p* < 0.1 in the univariate model were further entered into the multivariate analysis. All statistical testing was performed using 2-tailed tests; *p* < 0.05 was considered statistically significant. All analyses were performed using SPSS statistical software, version 22 (SPSS, Chicago, IL).

***CRISPR-DNMT1-interacting RNA (CRISPR-DiR): In vitro generation of sgRNA transcripts***

Approximately 1.4kb of the genomic fragment spanning the SALL4 5’ UTR‑exon 1‑intron 1 regionwas PCR amplified (Zymo Research) and cloned into the pGEM-T Easy vector. The vector was linearized with BamH1 restriction enzyme (New England Biolabs). SALL4-targeting sgRNA candidates were transcribed with HiScribe™ Quick T7 High Yield RNA Synthesis Kit (New England Biolabs) following the manufacturer’s instructions. The sgRNA target sequences within the SALL4 locus is 5’-CCAGGGCGAGCAGCAGCCGCA-3’, targeting the opposite strand and using CGG as the PAM sequence.

***CRISPR-DiR: In vitro cleavage and selection of sgRNA transcripts***

An in vitro cleavage assay was performed using purified Cas9 nuclease from *S. pyogenes* (New England Biolabs) in order to select SALL4-specific sgRNAs among a number of candidates. The experiment was performed according to the manufacturer’s protocol. The sgRNAs were denatured at 95⁰C for 3 minutes, then Cas9 protein and sgRNAs were incubated for 10 minutes at 25⁰C to form a complex. Lastly, a linearized SALL4 DNA fragment was added to the mixture and the entire reaction was incubated at 37°C for 1 hour. The reaction mixture was composed of purified Cas9 protein, an individual sgRNA, and a linearized SALL4 genomic fragment in a ratio of 10: 10: 1. 1 ul of Proteinase K was added to each sample after the cleavage reaction, and it was then incubated at room temperature for 10 minutes. The result was analyzed with a 1% agarose gel.

***CRISPR-DiR: Lentiviral transduction of DiR-SALL4 and dCas9***

Lentiviruses expressing dCas9 or sgRNA were packaged in 293T cells with the plasmids psPAX2 and pMD2.G. TransIT-LT1 Transfection Reagent (Mirus) was used for transfection into 293T cells. Virus was collected at 48 hours and 72 hours post-transfection. The collected virus was filtered through 0.45 µm microfilters and stored at -80 °C. Transduction of SNU-387 cells was performed by mixing virus and 4 μg/mL polybrene (Santa Cruz) together to add to the cells seeded in T75 flasks 24 hours prior to the transduction. 24 hours after the transduction, the medium was replenished with normal RPMI culture medium. Transduction efficiency was determined by GFP (for sgRNA) or mCherry (dCas9) expression by FACS analysis, and the positive cells were sorted by a FACS Aria machine (BD Biosciences).

***Real-time quantitative reverse transcriptase polymerase chain reaction (qRT-PCR)***

Total RNA was extracted with Trizol reagent (Invitrogen) according to the manufacturer’s instructions and treated with DNase. The RNA concentration was measured with ultraviolet spectrophotometry. Reverse transcription and PCR were performed using the iScript One-Step RT-PCR Kit with SYBR Green (Bio-Rad, Hercules, CA, USA; catalog no. 170-8893). Triplicate reactions were run for each gene. The expression level was normalized to glyceraldehyde 3-phosphate dehydrogenase (GAPDH). For each sample, an amplification plot and corresponding dissociation curves were examined. Relative quantification analysis was performed using the comparative CT method (2^−ΔΔCT^). SALL4 upregulation (SALL4^up^) and downregulation (SALL4^down^) between diagnosis and completion of 4 cycles of AZA treatment (t0 versus t4) were defined as 2-fold. The formula used to determine fold change is as follows: 2^−ΔΔCT^ = 2-^[ t4 CT (SALL4 after AZA) – t4 CT (GAPDH) ] –[ t0 CT (SALL4 at diagnosis)–t0 CT (GAPDH)]^, a scale of 1 to infinity and a scale of less than -1, were used to define SALL4^up^ or SALL4^down^, respectively. The sequences of primers for genes tested as follows: *SALL4* (*SALL4* exon3/4 span), forward primer 5′-AAGGCAACTTAAAGGTTCACTACA-3′, reverse primer 5′-GATGGCCAACTTCCTTCCA-3′; *SALL4A*-specific, forward primer 5′‑TGATCCCAACGAATGTCTCA-3′, reverse primer 5′-CCCAAGGTGTGTCTTCAGGT-3′; *SALL4B*-specific, forward primer 5′‑AAGCACAAGTGTCGGAGCA-3′, reverse primer 5′‑GTGCAGCCATGTTGCTTG-3′; *GAPDH*, forward primer 5′‑GAAGGTGAAGGTCGGAGTCAAC-3′, reverse primer 5′‑TGGAAGATGGTGATGGGATTTC‑3′.

***Western blot***

Western blot was performed according to standard protocols. The following antibodies were used for western blotting: SALL4 (ab29112, Abcam), and GAPDH (Santa Cruz, sc-25778). The dilution ratio of SALL4 antibody was 1:1000.

***Droplet digital polymerase chain reaction (ddPCR)***

Reactions for the ddPCR were prepared by harvesting 100,000 cells on each day for RNA extraction and cDNA preparation. The reaction mixture was prepared with the 2x ddPCRsupermix for probes (Biorad, Cat #186-3026), 10-fold diluted cDNA, nuclease-free water, and forward and reverse primers. Once the reaction mixture was ready, it was loaded onto the DG8 cartridge for the QX200 Automated Droplet Generator (Biorad, catalog no.186-4003). Thermal cycling was performed using the Biorad C1000 Touch Thermal Cycler with the following cycle conditions: 95°C for 10minutes, 94°C for 30 seconds (40 cycles), 60°C for 2 minutes (40 cycles), 98°C for 10 minutes, and 4°C hold. The reaction plate was loaded into the QX200 Droplet Reader (Biorad, Cat#186-4003) for gene expression analysis. To detect SALL4A and SALL4B, the following primers were used: SALL4A, forward primer 5' TGATCCCAACGAATGTCTCA‑3', reverse primer 5'‑CCTGAAGATGCATTATCGCA‑3'; SALL4B, forward primer5'‑GGTGGATGTCAAACCCAAAG‑3', reverse primer 5'-TCTCCCTTCCACGTTTATCC‑3'.

***Next generation sequencing (NGS) pipeline and validation methods***

DNA samples were extracted using the QIAamp DNA Mini Kit (Qiagen AG, Milan, Italy), in accordance with the manufacturer’s instructions. NGS data about patients used in this study were extrapolated from our main cohort of genetically screened MDS patients. DNA samples collected at the time of diagnosis were processed and analyzed as previously reported.^1^In brief, NGS screening for common somatic mutations in thirty genes known to be involved in MDS pathogenesis was performed according to the commercial Myeloid Solution by SOPHiA GENETICS (SOPHiA GENETICS, Saint-Sulpice, Switzerland) on a MiniSeq^®^ sequencing platform (Illumina, San Diego, California). The NGS analysis was performed on generated FASTQ sequencing files using the SOPHiA DDM^®^ platform that allows for detection, annotation, and pre-classification of genomic mutations (SNVs and Indels) through its SOPHiA™ artificial intelligence. Reads were aligned to the human reference genome (hg19 assembly). Only mutations with a VAF ≥ 1% (variant allele frequency), threshold coverage ≥ 1000x, and identified as highly or potentially pathogenic by the SOPHiA DDM^®^ platform were considered for all subsequent steps of the analysis. Single nucleotide polymorphisms (SNP), variants localized in the intronic and UTR regions, as well as synonymous variants were also excluded from the analysis. Targeted-NGS sequencing data are stored athttps://www.sophiagenetics.com (SOPHiA DDM platform), and can be extracted using the Sophia-DDM-v4 password-protected software. Raw data will be provided to researchers upon request. Validation of identified variants was performed using pyrosequencing technology (VAF ≥ 10%) and Sanger sequencing (VAF ≥ 20%). Pyrosequencing reagents (PyroMark Gold Q96, QiagenSrl, Milan, Italy), instrumentation, and software used for pyrosequencing analysis were as recommended by the manufacturers (PyroMark Q96 ID, DiatechPharmacogenetics,Jesi, Italy, PyroMark Assay Design and PyroMark Q24 version 2.0.6). Sanger sequencing reagents (BigDye Terminator v.3.1 cycle sequencing kit, Applied Biosystems/Life Technologies, Milan, Italy) and instrumentation were used for Sanger sequencing (ABI PRISM 3100; Applied Biosystems/Life Technologies, Milan, Italy). All primers were homemade designed, and all mutations were confirmed and quantified in independent experiments.

**Supplementary Figure Legends**

**Figure S1**. (A) SALL4 expression in 37 MDS patients at diagnosis before treatment in comparison to the controls (normal CD34(-) and CD34(+) cells). CD34‑ and CD34+: normal bone marrow; MDS: bone marrow mononuclear cells; CR: complete remission; PR: partial remission; HI: hematologic improvement (HI); SD: non-responders with stable disease; PD: progressive disease; (B) Log_2_ fold change of SALL4 based on responders and non-responders in 25 patients.

**Figure S2.** Survival based on the change of SALL4 expression and clinical response. (A) Progression˗free survival (PFS) between SALL4^up^ and SALL4^down^; (B) PFS based on treatment response and SALL4 expression change; (C) OS based on treatment response and SALL4 expression change.

**Figure S3. Distribution, frequency, and variant allele fraction (VAF) of somatic mutations in 25 MDS patients.** Light-, intermediate-, and dark- black boxes indicate the presence of 1, 2, or ≥2 mutations in the same gene, whereas empty boxes indicate wild-type genes.

**Supplementary Figures**

**Figure S1**

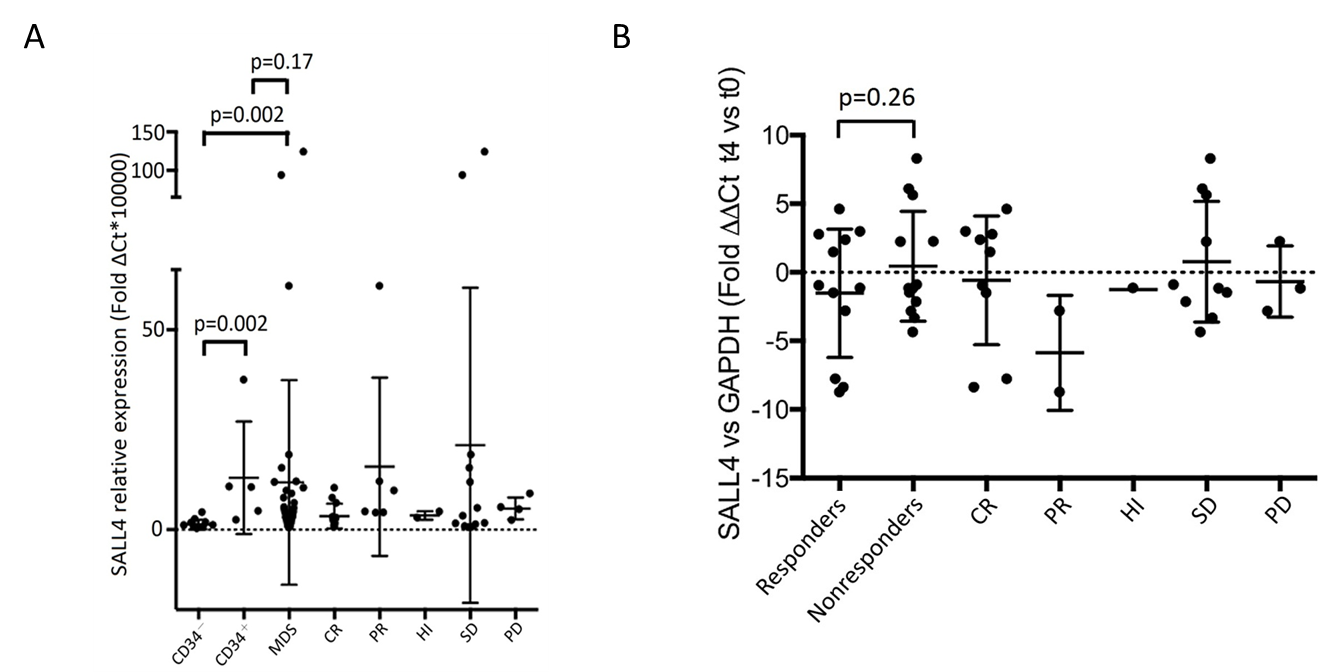

**Figure S2**

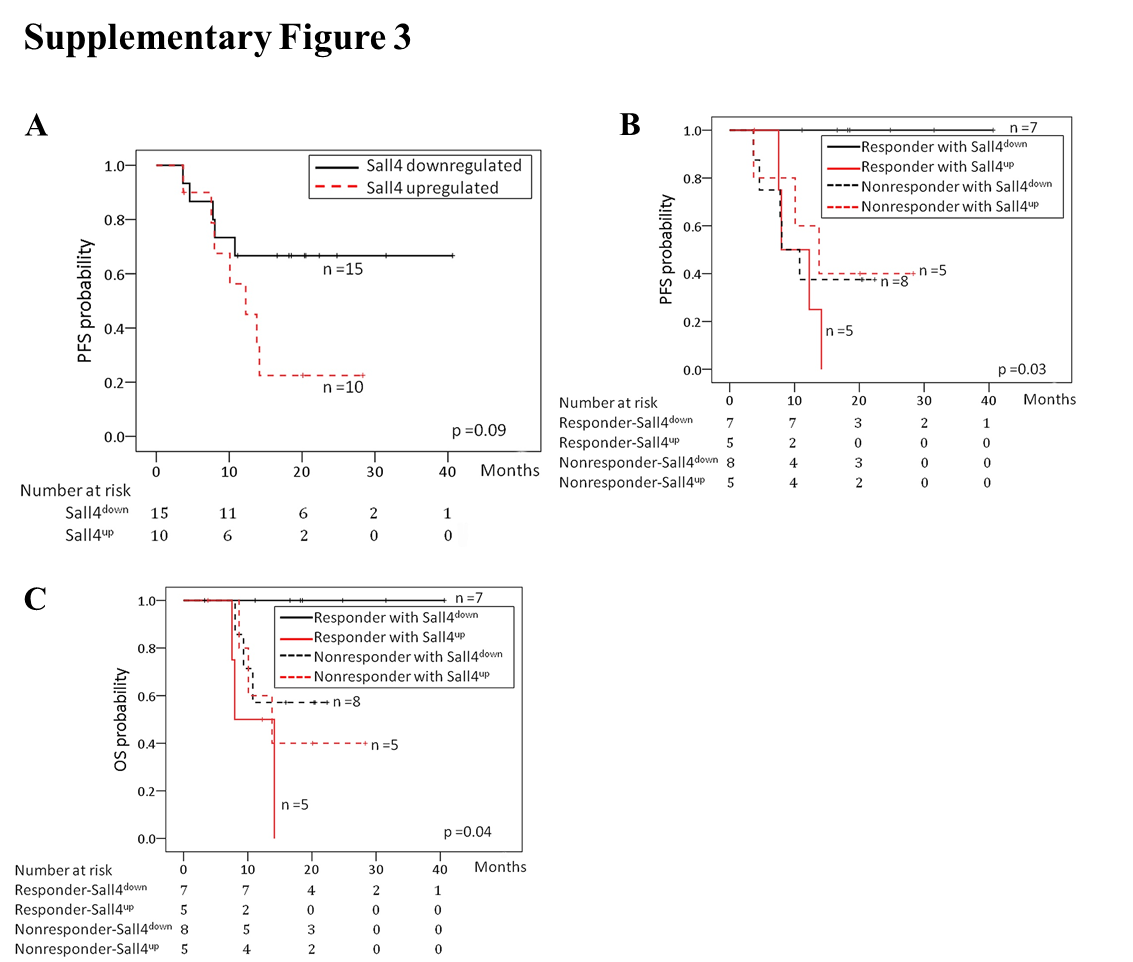

**Figure S3**

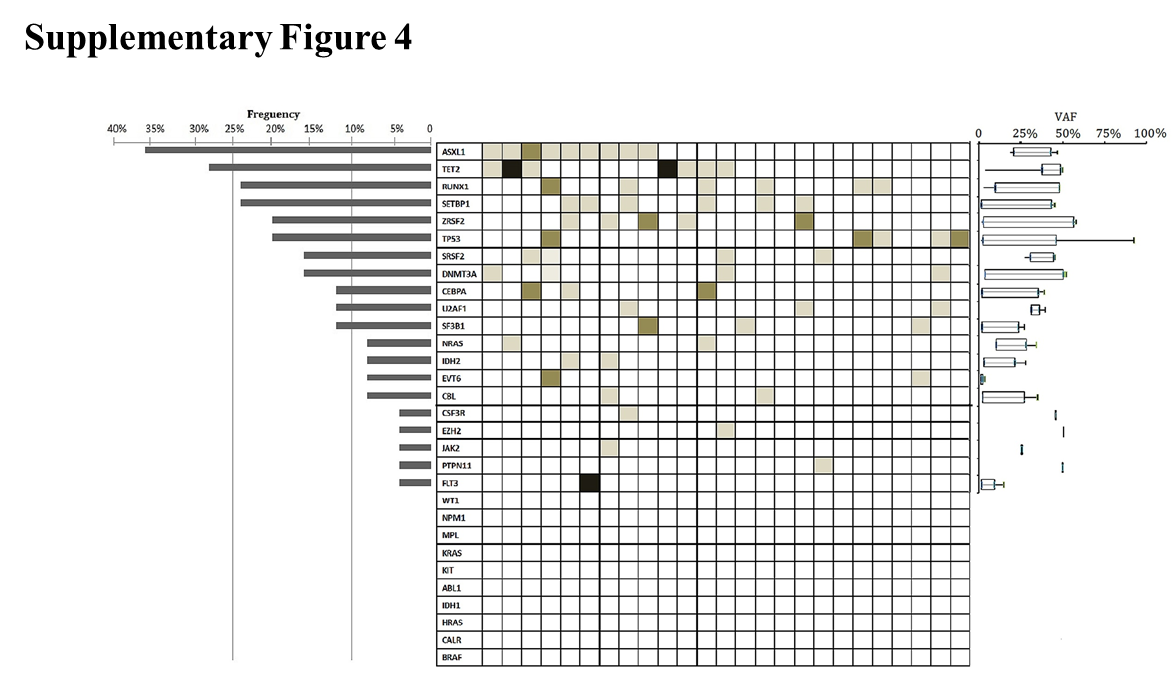

**Supplementary table**

| **Table S1. Prognostic factors of twenty-five MDS patients in OS** | | | | | | | |
| --- | --- | --- | --- | --- | --- | --- | --- |
|  | Univariate analysis | | |  | Multivariate analysis | | |
| Parameters | HR | 95% CI | *p*-value |  | HR | 95% CI | *p*-value |
| Age ≥ 60 years | 1.84 | 0.49-6.92 | 0.36 |  |  |  |  |
| Male | 4.06 | 0.50-32.56 | 0.18 |  |  |  |  |
| Nonresponders | 1.86 | 0.46-7.47 | 0.38 |  |  |  |  |
| SALL4 upregulation after AZA. | 4.24 | 1.05-17.22 | 0.03 |  | 6.48 | 1.06-39.67 | 0.04 |
| IPSS intermediate-2 or high risk | 24.71 | 0.00-x | 0.49 |  |  |  |  |
| ANC < 1500/µl | 0.42 | 0.10-1.72 | 0.23 |  |  |  |  |
| Hemoglobin < 10g/dL | 2.04 | 0.50-8.25 | 0.31 |  |  |  |  |
| Platelet count < 100000/µl | 0.93 | 0.23-3.74 | 0.92 |  |  |  |  |
| Poor karyotype | 2.81 | 0.66-11.89 | 0.15 |  |  |  |  |
| Mutational profiles* |  |  |  |  |  |  |  |
| ASXL1 mutation | 1.97 | 0.52-7.40 | 0.31 |  |  |  |  |
| TET2 mutation | 0.21 | 0.02-1.71 | 0.14 |  |  |  |  |
| RUNX1 mutation | 3.91 | 1.00-15.19 | 0.04 |  | 10.66 | 1.25-90.72 | 0.03 |
| SETBP1 mutation | 4.45 | 1.16-17.04 | 0.02 |  | 1.85 | 0.37-9.12 | 0.44 |
| TP53 mutation | 2.80 | 0.68-11.46 | 0.15 |  |  |  |  |
| ZRSF2 mutation | 3.64 | 0.85-15.52 | 0.08 |  | 2.94 | 0.30-28.19 | 0.34 |
| DNMT3A mutation | 0.70 | 0.08-5.65 | 0.74 |  |  |  |  |
| SRSF2 mutation | 0.48 | 0.06-3.85 | 0.49 |  |  |  |  |
| HR, Hazard ratio; CI, Confidence Interval; SD, Stable disease; PD, Progression disease; AZA, Azacytidine; SALL4, Spalt like transcription factor 4; IPSS, International Prognostic Scoring System; ANC, Absolute neutrophil count  x:269250.47  *Mutations present in less than four patients were excluded from the analysis | | | | | | | |
